# Automatic radiotherapy treatment planning with deep functional reinforcement learning

**DOI:** 10.1101/2024.06.23.24309060

**Authors:** Bin Liu, Yu Liu, Zhiqian Li, Jianghong Xiao, Guosheng Yin, Huazhen Lin

**Affiliations:** Center of Statistical Research, School of Statistics, Southwestern University of Finance and Economics, Chengdu, China; Radiotherapy Physics & Technology Center, Cancer Center, West China Hospital, Sichuan University, Chengdu, Sichuan, China; Department of Statistics and Actuarial Science, The University of Hong Kong, Hong Kong, China

## Abstract

**Background and purpose:** Intensity-modulated radiation therapy (IMRT) is a crucial radiotherapy technique, often formulated as an optimization problem. However, when the constraints are too tight to provide a feasible solution, human planners resort to relaxing the optimization parameters and re-evaluating until an acceptable solution is obtained. This process is laborious and time-consuming which has prompted attempts to automate radiotherapy through inverse planning studies using reinforcement learning. Unfortunately, these studies face two major limitations. Firstly, a separate sub-network must be designed for each organ, rendering them difficult to apply to patients with an inconsistent number of structures. Secondly, the low signal-to-noise inputs and discrete action space result in low training efficiency. To address these limitations, this study proposes a novel and effective model.

**Methods:** This study proposes an organ-sharing network called *F*unctional *a*utomatic *t*reatment *P*lann*I* ng *N*etwork (FatPIN), which contains a functional embedding layer to extract curve features of the dose-volume histogram (DVH). It outputs continuous actions that adjust the optimization parameters, thereby automating the radiotherapy planning process.

**Results:** Experiments were conducted on the cervical cancer dataset and the results show that the FatPIN is feasible and effective in real-world radiotherapy. With automatic iteration, FatPIN gradually increased the PTV dose, while reducing the dose levels of the OARs. Specifically, at step 50, the *D*_95_ of the PTV reached 51.68 Gy, exceeding the clinical standard of 50.40 Gy, the *V*_30_, *V*_40_ and *V*_50_ of all OARs were within clinical requirements.

**Conclusion:** We proposed FatPIN to implement automatic radiotherapy treatment planning. Experimental assessments conducted on cases of cervical cancer demonstrate significant improvements in patient metrics facilitated by FatPIN, thus confirming its practical applicability in clinical contexts.

## 1 Introduction

Cancer is a leading cause of death worldwide, accounting for approximately 10 million deaths in 2020 (nearly one in six deaths worldwide) [1]. Among the various treatment modalities available, radiation therapy plays a pivotal role in effectively managing cancer. It utilizes concentrated beams of high-energy radiation to target and control the planning target volume(s) (PTVs). However, a significant challenge arises due to the presence of organs-at-risk (OARs) in close proximity to the targeted area. The beams employed in radiation therapy not only aim to eradicate cancer cells but also inadvertently affect adjacent healthy tissues, which comprise the OARs. To address this challenge, inverse treatment planning techniques, such as intensity-modulated radiation therapy (IMRT), have emerged as crucial components of modern radiation therapy strategies.

In practice, achieving IMRT involves a complex optimization process as shown in eq. (12). The objective of the optimization typically contains several dose-level constrains to maximize the delivery of radiation of targets and minimize the hurts for OARs simultaneously. However, the formulation of the optimization problem of IMRT is typically based on a set of empirical parameters, such as relative importance weights and constraint levels for PTVs and OARs, which aim to accommodate various clinical considerations. These empirical parameters are initially initialized by human planners but can be adjusted as needed. To determine the optimal solution for a given set of parameters, a modern treatment planning system (TPS) is employed. However, it is not uncommon for the resulting solution to be inconsistent with the desired clinical considerations. In such cases, the planner must manually adjust these parameters and re-optimize the treatment plan using the TPS. This trial-and-error process is often repeated multiple times until a satisfactory result is achieved. Furthermore, the quality of the resulting treatment plan heavily relies on the experience and expertise of the planners. Consequently, there is a significant need to develop automatic methods that can produce high-quality and efficient treatment plans, thereby reducing the reliance on manual intervention.

Over the past years, significant efforts have been made to develop automatic planning methods. A typical solution is to add an outer-loop optimization for parameter searching on top of the inner optimization. Several notable examples of this double-optimization approach can be found in [2, 3, 4]. More recently, attention has turned towards the potential of deep learning in automatic treatment planning [5]. Deep neural networks offer greater flexibility in the parameter searching compared to traditional methods. Particularly, the trial-and-error process of human planners can be effectively emulated using deep reinforcement learning techniques. Indeed, studies have demonstrated that deep reinforcement learning can be employed to adjust treatment plan parameters[6, 7, 8].

For instance, Shen et al.[9] propose a deep reinforcement learning-based virtual therapy planner network to model the decision-making behavior of human planners when adjusting treatment planning parameters (TPPs) for prostate cancer. However, existing methods suffer from several limitations, 1) Firstly, they typically design separate sub-networks for each organ [10, 7], making it challenging to generalize their models to different cases. This is because the number of organs involved in treatment planning can vary among patients, even for those with the same disease. 2) Secondly, these methods neglect the functional characteristics of the dose-volume histogram (DVH) curve and often take it as an image, resulting in a significant reduction in the signal-to-noise ratio of the input. Consequently, the learning process becomes more challenging, and the number of organs that can be effectively planned is limited. For example, as discussed in [6, 10], these methods only consider the bladder and rectum as OARs, while clinical settings frequently involve adjusting more than a dozen organs. 3) Thirdly, existing methods model the process of dose adjustment as a classification problem, for example, setting five possible TPPs adjustment options {+2%, +1%, 0, -1%, -2%}, corresponds to increase or decrease TPP values by 2% or 1%, or keep them unchanged [9]. However, this classification approach severely restricts the search capability of the model. Therefore, there is substantial potential for advancing the generalization of multi-parameter TPP adjustment across multiple organs.

In this study, we introduce a deep reinforcement learning framework that takes into account the functional characteristics of the dose-volume histogram (DVH). We refer to this framework as the *F*unctional *a*utomatic *t*reatment *P*lann*I* ng *N*etwork (FatPIN). The primary objective of FatPIN is to train a model capable of manipulating the TPS and adjusting treatment planning parameters to generate high-quality treatment plans automatically. This automated approach aims to enhance the efficiency and accuracy of treatment planning by leveraging the capabilities of deep reinforcement learning. The ultimate goal is to deliver treatment plans that effectively adhere to clinical standards while reducing the reliance on manual intervention.

In the realm of clinical practice, the condition of patients displays notable variability. Even among individuals diagnosed with the same type of cancer, the specific considerations for radiation therapy by medical practitioners may diverge due to unique factors such as tumor stage and organ size. The presence of these individualized requirements presents challenges when implementing a model predicated on a singular organ corresponding to a sub-network in a clinical context.

To mitigate this issue, FatPIN incorporates a neural network architecture tuned for organ-sharing. This innovative design facilitates the incorporation of DVHs from all tissue structures (organs) into the same network during the training phase. Subsequently, in testing, the DVHs of diverse structures for a particular patient can be directly input into the network to yield adjusted action prediction results. Essentially, this approach removes the constraint on the number of organs involved in treatment planning imposed by the model.

To bolster the learning efficiency of FatPIN, a functional decomposition layer is employed to initially extract features from the DVH curves. This functional mapping layer markedly enhances the signal-to-noise ratio of the input data. Additionally, departing from the conventional method of modeling dose adjustment as a classification problem, we propose learning the distribution of dose adjustment. Specifically, we establish a connection between the output of FatPIN and the mean and variance of the dose adjustment distribution. Ultimately, parameterization techniques are utilized to generate continuous dose adjustments based on the acquired mean and variance. This strategic approach enhances the overall efficiency of model training, facilitating its application in real-world scenarios with heightened effectiveness and reliability.

## 2 Continuous Functional Treatment Parameters Adjustment Networks

Manual adjustment of dosage and weight to achieve a high-quality treatment plan is a cumbersome and time-consuming process in clinical radiotherapy. Therefore, automatic radiotherapy regimens are required with automatic dose and weight adjustments by planners. The overall framework will be introduced in next section.

### 2.1 Overall Procedure

Figure 1 depicts the overall framework of our proposed method, *F*unctional *a*utomatic *t*reatment *P*lann*I* ng *N*etwork (FatPIN). It comprises three essential modules: a functional embedding layer, a continuous treatment planning neural network, and an environment for deep reinforcement learning (DRL). The overall process of automatic planning can be divided into the following steps:

**Figure 1:**
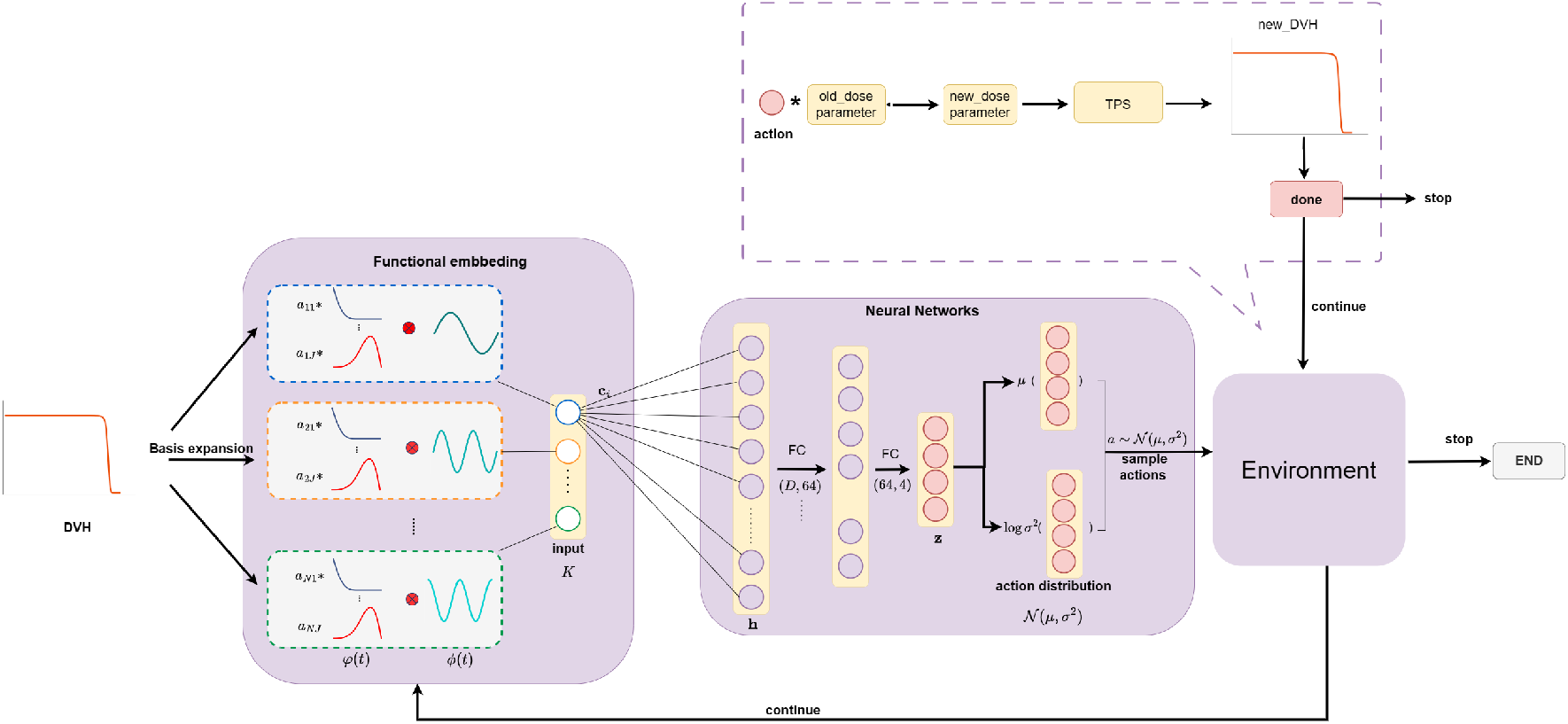
The overall framework of the proposed FatPIN.

1. The functional embedding layer accepts DVHs (state) *s*_*t*_ = (*s*_*t*_(1), *s*_*t*_(2), …, *s*_*t*_(200)) at time *t* as input and outputs the functional embedding of DVHs *x*_*t*_.
2. The treatment planning neural network take functional embedding of DVH ***x***_*t*_ as input and yield a compressed feature **z**_*t*_;
3. Estimates the mean ***µ***_*t*_ ∈ ℝ^*M*^ and the logarithmic form of variance 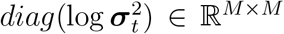 of the TPP action ***a***_*t*_ ∈ ℝ^*M*^ for the *M* tissue structures with **z**_*t*_;
4. Randomly samples TPP actions: 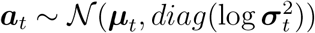;
5. Updates the constraints of the optimization with ***a***_*t*_ (see eq. (15)).
6. Solves the new optimization problem with TPS and yields a corresponding new state ***s***_*t*+1_ (updated DVH) and calculates the corresponding reward *r*_*t*_;
7. Stores (***a***_*t*_, ***s***_*t*_, *r*_*t*_, ***s***_*t*+1_) into sample set *C*;
8. Updates the parameters of FatPIN and repeats the above steps until convergence.

The following sections will introduce the details of the modules involved in the above process.

### 2.2 Functional Embedding Layer

#### 2.2.1 Dose-volume Histogram

In radiotherapy planning, dose-volume histograms (DVH) ***s*** is a curve that depicts the distribution of the dose for each structure present in the treatment area, including the OAR and the target. Figure 2 presents a typical example of a DVH for an OAR and a target in a treatment plan. The horizontal axis in the DVH represents the dose level, while the vertical axis represents the fraction of the volume. For instance, the point P in the target DVH in Figure 2 indicates that at least 60% of the volume of the target voxels receive a dose level of 1.8 Gy or less. Ideally, the DVH in the target area should be vertical at the prescribed dose, indicating that all the voxels in PTV receive the prescribed dose, while the OAR should be vertical at a relative dose of 0, indicating that it receives a dose of 0. Oncologists may be willing to sacrifice a certain portion of the OAR close to the target area to ensure optimal tumor control probability. Thus, OAR must be required to have at least a certain percentage of the dose below the specified level to reduce the risk of damaging the nearby healthy tissue.

**Figure 2:**
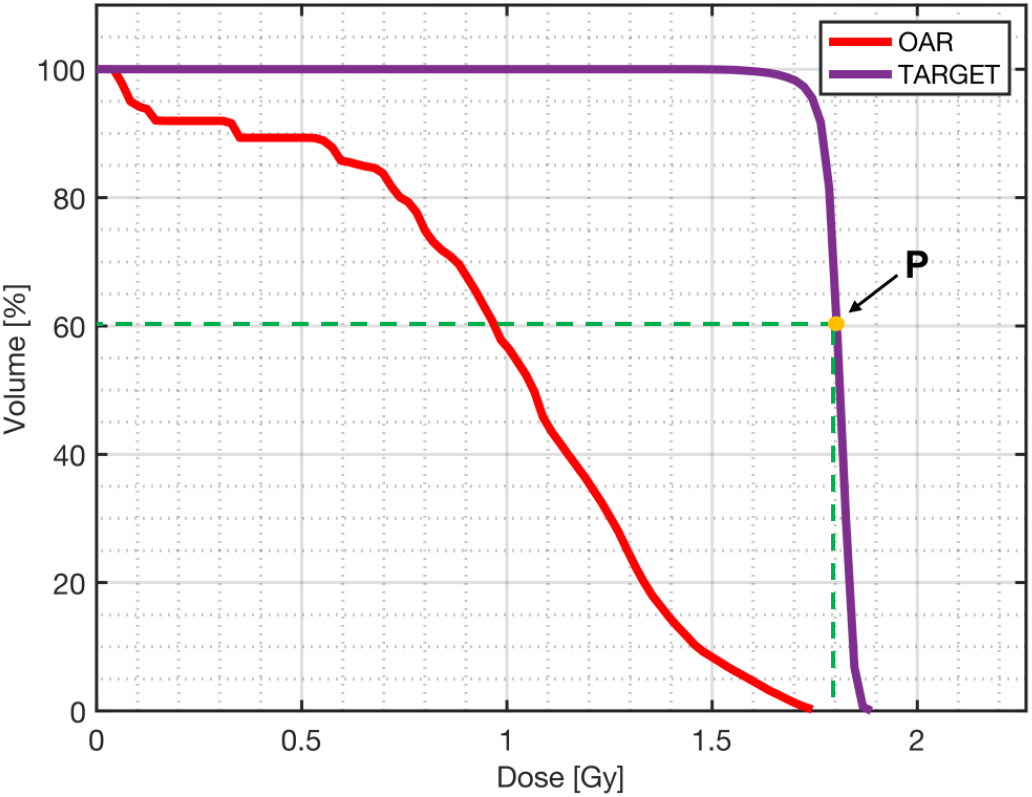
Example of DVH curves for an OAR and a target. The DVH observations *s*_*t*_(*u*) ∈ [0, 1] correspond to a functional relationship between the dose level *u* [Gy] (or relative dose level) of the horizontal axis and the fraction of volume *s*_*t*_(*u*) receiving that dose level of the vertical axis. This functional relationship provides a comprehensive representation of the radiation dose distribution within the target and the OARs.

#### 2.2.2 Functional Embedding of DVH

Instead of directly feeding the DVH curves into the DRL model, FatPIN embeds them into finite-dimensional vectors. We assert that these embedding vectors are a more suitable input for subsequent adaptation to standard deep neural networks. The details are as following.

Let *s*_*t*_(*u*) denotes a single input DVH curve at timestamp *t* with horizontal axis *u*, the calculation of a hidden neuron in the neural network is

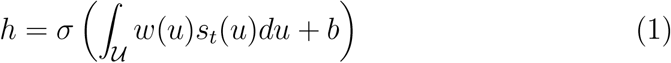

where *w*(*u*) are functional weights and *b* is the intercept term. We further decompose *w*(*u*) using its basis representation,

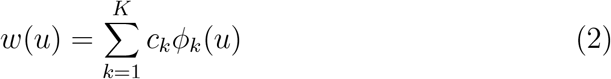

where *ϕ*_*k*_(*u*) represents the basis function of the functional weights *w*(*u*). This basis function can be a Fourier basis or a spline basis, and we choose *K* as the number of basis functions. In our approach, we select *ϕ*_*k*_(*u*) to be the Fourier basis,

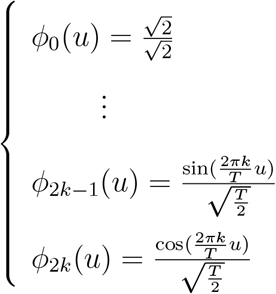

and set *K* = 5 as shown in Figure 3.

**Figure 3:**
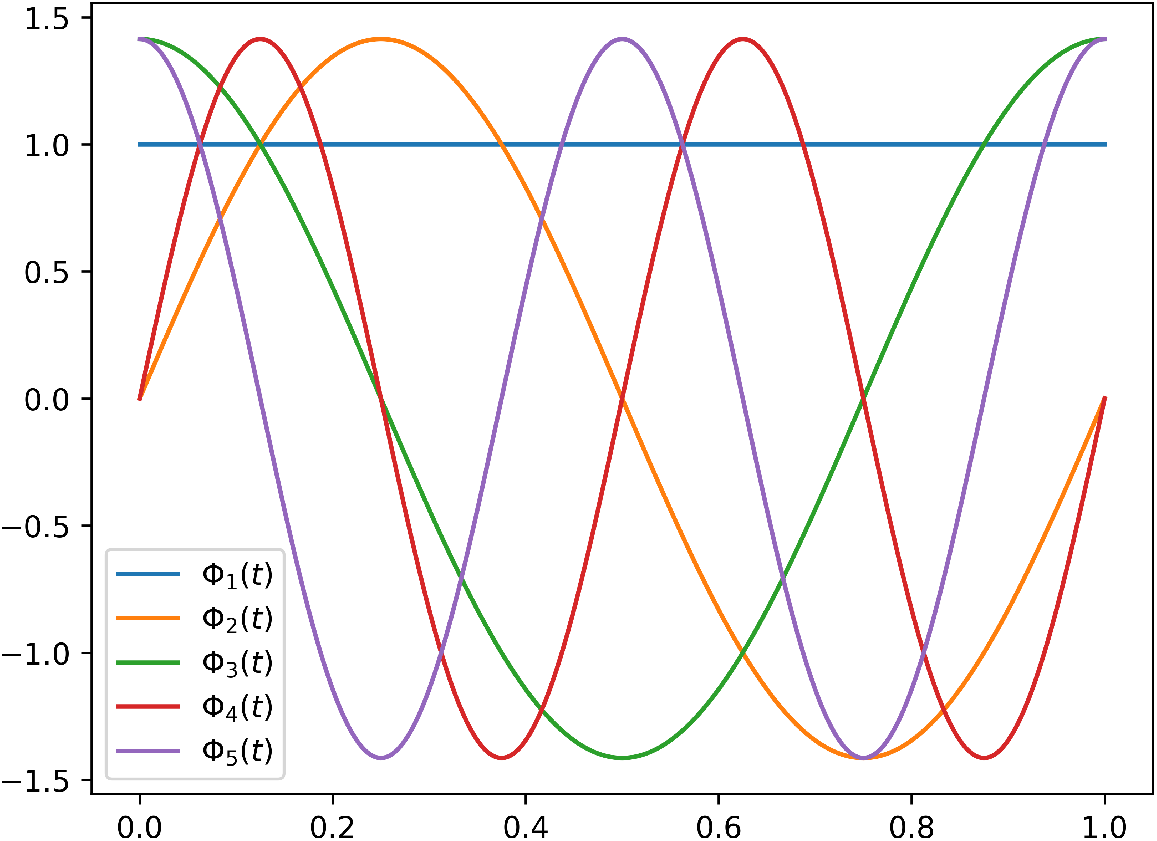
The 5 Fourier basis *ϕ*(*u*).

Therefore, we can reformulate eq. (1) as,

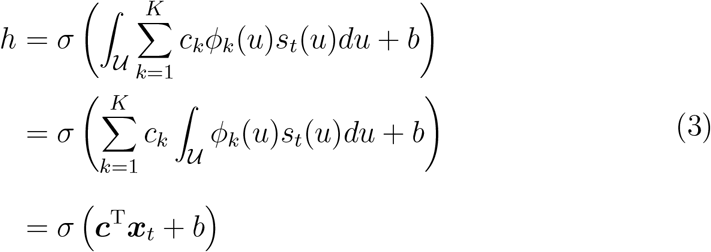

where

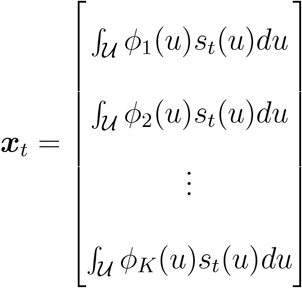

is the functional embedding of DVH what we try to get. In practice, the integral ∫_𝒰_ *ϕ*_*k*_(*u*)*s*_*t*_(*u*)*du* in eq. (3) can be approximated with numerical integration methods such as the composite Simpson’s rule.

As for effectively handling the DVH curve *s*_*t*_(*u*), we employ a B-spline basis expansion approach. That is, we express each DVH as a linear combination of *J* B-spline bases, which can be represented as follows:

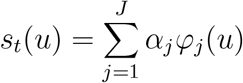

where *α*_*j*_, *j* = 1, 2, …, *J*, is the corresponding coefficients of *s*_*t*_(*u*) with respect to the basis *φ*_*j*_ (*u*), *φ*_*j*_ (*u*) is the B-spline basis, *J* is the number of the B-spline basis, we set *J* = 35 as shown in fig. 4.

**Figure 4:**
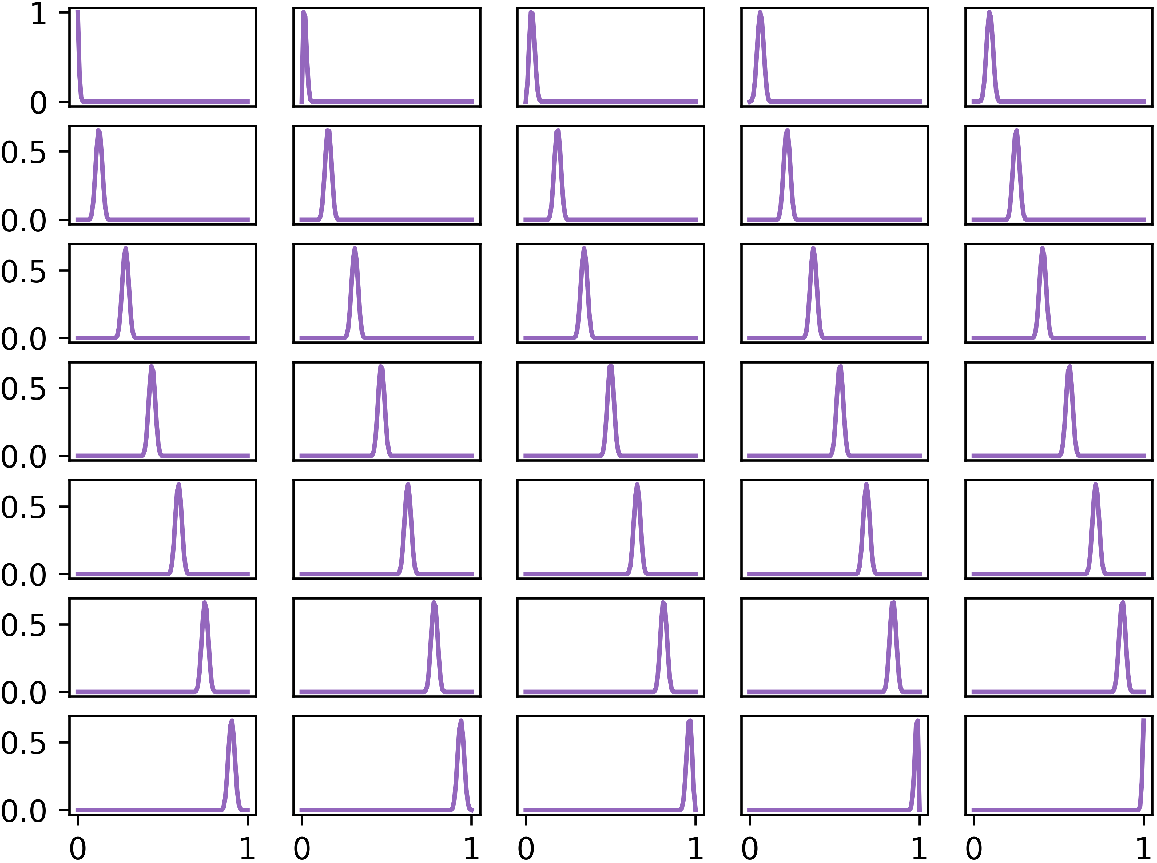
The 35 B-spline basis *φ*(*u*).

In our problem, we often encounter scenarios where multiple organs are involved, resulting in a set of input DVHs. Let’s assume that *M* is the number of organs that need to be adjusted, then we will get functional embedding of DVHs *X*_*t*_ ∈ *R*^*M*×*K*^ in matrix form.

### 2.3 Treatment Planner Neural Networks

We use a similar framework of Deep Deterministic Policy Gradient [11] to implement the virtual treatment planner neural networks. The planner network can output actions ***a*** ∈ ℝ^*M*^ for *M* structures. Suppose *a* is *i*-th component of ***a***, which corresponds to the action of the *i*-th tissue structure. The process of estimate *a* with the virtual treatment planner neural networks is as follows.

In the context of deep reinforcement learning, our virtual treatment planner neural network acts as an agent that aims to interact with the environment. This network comprises two parts: the actor network *m* (***s***|*Θ*) is tasked with learning a policy for the agent and the critic network *Q* (***s***, *a*|*Φ*) aims to estimate the action-value function *Q*_*π*_ (***s***_*t*_, *a*_*t*_).

To fully explore the environment, we add two linear layers to respectively output the mean *µ*_*t*_ and the logarithmic form of variance 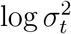 of action *a*_*t*_ based on the original actor network output **z**_*t*_ as follows,

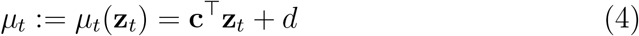

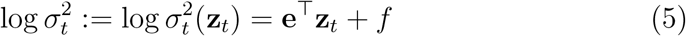

where **c**, *d*, **e**, *f* are parameters of the linear layers. Then, we use them to generate a new action *a*_*t*_ with

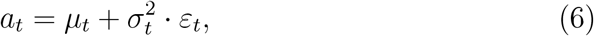

where *ε*_*t*_ ∼ 𝒩 (0, 1). The action *a*_*t*_ is exactly what our new actor network outputs.

To make the critic network’s action estimate as close as possible to the reward-based action estimate, the loss function of original critic network *Q* (***s***, *a*|*Φ*) is

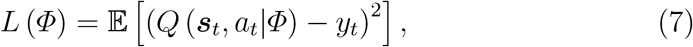

where

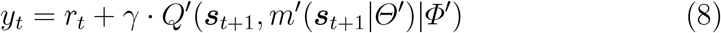

is TD (Temporal Difference) target [12], *γ* is a discount parameter, *m*^′^(***s***|*Θ*^′^) and *Q*^′^ (***s***, *a*|*Φ*^′^) are two target networks, which are copies of the learned networks *m* (***s***|*Θ*) and *Q* (***s***, *a*|*Φ*). We update the target networks in a soft way, that is, *θ*^′^ ← *δθ*+(1 − *δ*) *θ*^′^ with *δ* ≪ 1, where *θ* and *θ*^′^ are the parameters corresponding to the learned network and the target network respectively.

Based on the preceding statement, we hope to make the optimization of the action *a* more precise, so it is necessary to control the magnitude of its variance 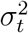. Therefore, we have added a penalty term for the variance to regularize the loss function as following,

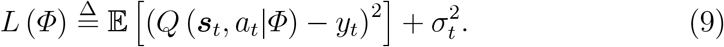

### 2.4 Model Training

The DVH ***s***_*t*_(*u*) is a curve that can be represented by coordinates. In this work, we record the DVH using 100 2-D points. Therefore, the curve ***s***_*t*_(*u*) can be expressed as a 200-dimensional vector ***s***_*t*_(*u*) = (*s*_*t*1_, *s*_*t*2_, …, *s*_*t*200_), where the first 100 dimensions correspond to the horizontal coordinates, and the subsequent 100 components represent the corresponding vertical ordinates.

At iteration step *t*, the functional embedding layer firstly accepts the ***s***_*t*_ (DVH of a structure at time *t*) as input and outputs its functional embedding ***x***_*t*_ (section 2.2). Next, we feed the functional embedding of DVH ***x***_*t*_ for the actor network *m* (***s***|*Θ*) to predict the mean *µ*_*t*_ and the logarithmic variance log 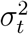 of the dose adjustment action *a*_*t*_. Then, we sample a random action *a*_*t*_ using eq. (6).

Then the agent interacts with the environment by updating the optimization problem with *a*_*t*_ (as shown in eq. (15)). Upon solving the new optimization problem, we can calculate the corresponding reward *r*_*t*_ and obtain the updated state ***s***_*t*+1_. The sequence of transitions (***s***_*t*_, *a*_*t*_, *r*_*t*_, ***s***_*t*+1_) is stored in a replay buffer *R*. During model training, a batch of transitions is randomly selected from *R* to calculate *Q*(***s***_*t*_, *a*_*t*_|*Φ*) using the critic network. Then the temporal difference (TD) target is computed as in eq. (8). With *y*_*t*_, the loss can be calculated using eq. (9). That is, we can update parameters of the critic network *Q* (***s***, *a*|*Φ*) with the TD algorithm [12]. For the actor network *m* (***s***|*Θ*),we calculate the gradient of objective function *J* (*Θ*) as follows,

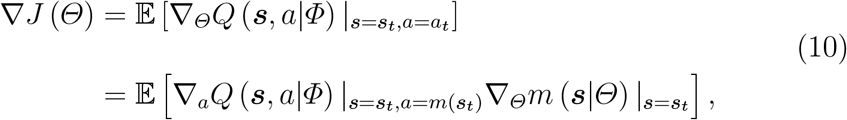

the parameters of the actor network can then be updated using policy gradient ascent. Finally, soft target updates are used to update the parameters of the two target network. We set training stop conditions based on clinical expertise. For example, the algorithm stops updating when the D95 in the target area exceeds its prescribed dose, and when the V50 of the OAR exceeds its prescribed dose.

### 2.5 Environment

The interactive environment for reinforcement learning, as depicted in Figure 1, receives adjustment actions ***a***_*t*_ derived from the agent (virtual treatment planner network) as input. These actions are utilized to create a new optimization problem by modifying its original constraints. The new optimization problem is then solved by the TPS, resulting in a new DVH. Rewards are determined by comparing the new DVH with the previous one. Further details regarding the reward computation can be available in section 2.6.

#### 2.5.1 Optimization Formulating

Suppose there are *M* structures that need to be planned, including the PTV and OARs. For our case, which involves cervical cancer, *M* = 12, and the OARs include the bladder, rectum, small intestine, bilateral femoral heads, and five ring structures.

Let the *m*-th organ be partitioned into *N*_*m*_ cubic volume elements or voxels of equal size [13](Typically greater than 2 mm in side length). Hence the dose for the *m*-th organ is represented by a vector 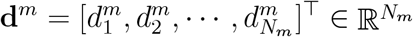, and Σ_*m*_ *N*_*m*_ = *N*, for *m* = 1, …, *M*, where *N* is the total number of voxels of the patient. Then the dose distribution for the *M* organs can be represented as **d** = [**d**^1^; **d**^2^; …; **d**^*M*^ ] ∈ ℝ^*N*^.

In order to achieve the required dose depicted by **d**, we will employ *L* beamlets at various angles to deliver radiation therapy to the *M* structures. Let Ω_*ij*_ ≥ 0 represent the dose deposition, which depicts the *i*-th voxel element by the *j*-th beamlet at unit beam intensity [13]. Ω ∈ ℝ^*N* ×*L*^ can be calculated by TPS, such as matRad [14]. It depends on the patient’s personal structure parameters.

Then the goal of IMRT is to find, for each beamlet and according to the optimization goals of the respective model, a suitable nonnegative beamlet weight defining its radiation intensity. Specifically, the dose *d*_*i*_ of voxel *i* can be computed as follows,

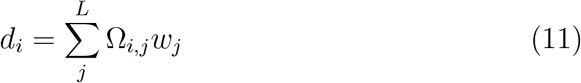

where *w*_*j*_ is the weight of beamlet *j*. The parameter *w*_*j*_ is the variable to be optimized. However, in this study, the optimization of *w*_*j*_ is indirectly achieved through the matRad optimization engine. In other words, we optimizes the dose vector **d** instead of *w*.

In the context of IMRT, the planning process involves solving a series of optimization problems that consist of well-defined objective functions and constraints. In our work, we adopt an overall optimization objective expressed as a weighted sum of various individual components as shown in eq. (12),

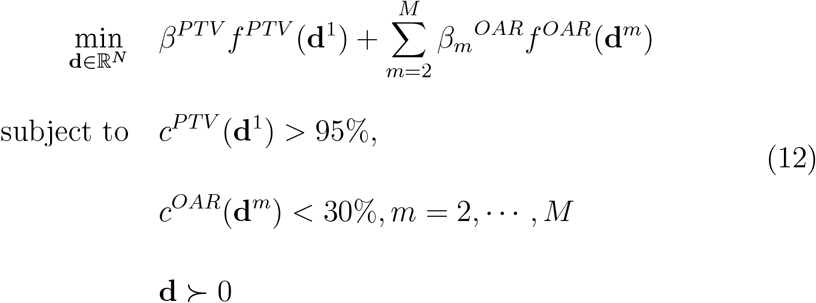

where *f* ^*PTV*^ (**d**^1^) is the objective function of PTV with a relative weighting (penalty) denoted as *β*^*PTV*^, and *f* ^*OAR*^(**d**^*m*^) denotes the objective function of *m*-th OAR with relative weighting 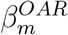. The corresponding constraint *c*^*PTV*^ (**d**^1^) *>* 95%ě indicates that 95% of the PTV voxels are required to receive a dose no lower than the prescription dose. The constraint *c*^*OAR*^(**d**^*m*^) *<* 30%ě indicates that no more than 30% OAR voxels are required to receive the prescription dose. The positivity constraint **d** ≻ 0 ensures that only positive radiation fluences are considered. In the implementation, we set *β*^*PTV*^ = 80 and 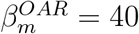.

In this work, *f* ^*PTV*^ (**d**^1^), *f* ^*OAR*^(**d**^*m*^), and the corresponding constraints are chosen as follows [14],

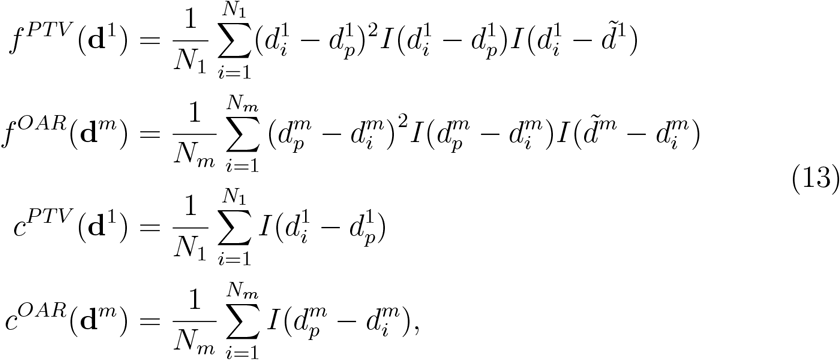

where *N*_1_ is the number of PTV voxels, *N*_*m*_ is the number of voxels of *m*-th OAR, 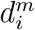 represents the dose in voxel *i* of the *m*-th structure, 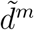 is prescribed volume, and 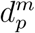 is the prescribed dose of *m*-th organ. *I*(*x*) is Heaviside function defined as,

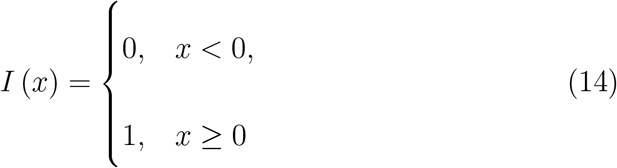

and 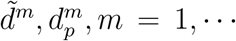, *M* are prescribed volume and prescribed dose for *m*-th structure [14]. Typically, 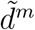 and 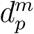 are parameters determined by the doctor. Different values for 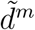 and 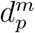 correspond to distinct constraints for the optimization problems in eq. (12). In our study, we fix 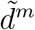 and adjust 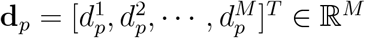 using the proposed method as follows,

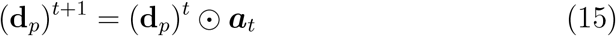

where ⊙ denotes the element-wise product. It suggests that action ***a***_*t*_ updates **d**_*p*_ and consequently gives rise to a new optimization problem as we stated in section 2.3. The updated objective function and constraint equations initiate a new round of optimization to obtain the results of this iteration as shown in Figure 1.

DVH is a criterion for evaluating the performance of the treatment plan. To drive the deep reinforcement learning process, we input an initial DVH into the ATPAN as a start. Instead of using the original DVH (an image that plotting the original DVH curves), we proposed to use its functional embedding as introduced in section 2.2. The input DVHs involve multiple organs. Suppose there are *N* organs require to adjust. A linear basis expansion is used with *i* basis functions defining each of these *N* DVHs. The embedding vectors of these DVHs are as defined follows,

### 2.6 Reward function

Based on the updated DVHs, we can compute the reward for the current iteration in the proposed model. The learning objective of our model is to maximize the dose delivered to the target while minimizing the dose received by the OARs. Consequently, we consider the change in the area between the DVH curves of the target *A*_*Target*_ and OARs *A*_*OARs*_ as a component of the reward. Additionally, the change in the D95 metric, which represents the dose received by 95% of the target volume, is also considered an important indicator that captures the attention of clinicians. Therefore, we incorporate the change in D95 as another source of reward. The final reward can be expressed as follows,

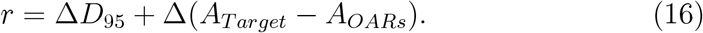

### 2.7 Cohort

In this study, a cohort of cervical cancer patients who underwent treatment with IMRT at our institution was selected. The treatment planning process involved defining the PTV as well as the organs at risk (OARs). The prescription dose for all patients was set at 50.40 Gy, delivered in fractions of 1.8 Gy each. The treatment plans for IMRT were optimized using the matRad software. Specifically, a configuration of equally spaced 5 coplanar photon beams was employed for all patients. These beam arrangements were then optimized using the IPOPT optimization algorithm within the matRad framework. The goal of the optimization process was to achieve an optimal dose distribution that effectively targeted the PTV while minimizing the dose to the surrounding OARs.

## 3 Results

### 3.1 Visualizations of Planning

Figure 5(a) and Figure 5(b) visualize the process of parameter adjustment. In these representations, the dash line corresponds to the Planning Target Volume (PTV), while the solid line represents the five Organs at Risk (OARs): Bladder, Femoral Head R/L, Rectum, and Small Intestine. As shown in Figure 5(a), during the initial five steps, FatPIN trys to reduce the dose levels of the five OARs while concurrently increasing the PTV dose to 50.4 Gy. We observe that the Small Intestine registers the lowest dose level, while the Bladder exhibits the highest dose level among the five OARs. The initial dose of the Small Intestine is approximately 45 Gy, decreasing to 20.42 Gy by the end of the iteration, aligning with the prescribed dose for OARs.

**Figure 5:**
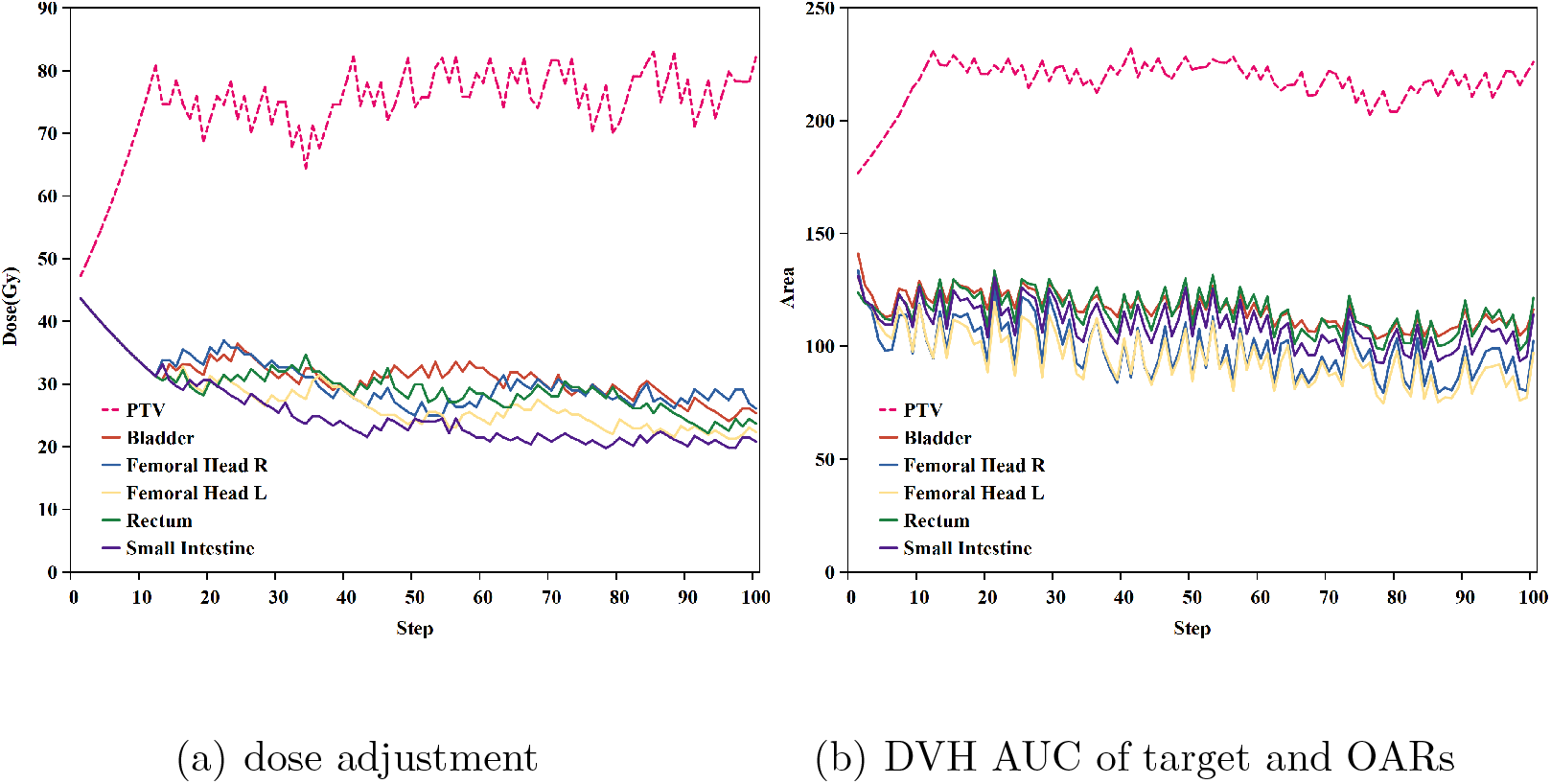
The process of parameter adjustment of FatPIN.

Commencing from the initial adjustment, FatPIN initiates the process of adjusting the PTV dose level, gradually increasing it while simultaneously reducing the dose level of the OARs until convergence is achieved around step 50. It is noteworthy that FatPIN employs an alternating approach in which the dose levels of both the PTV and OARs are adjusted. Figure 5(b) illustrates the changes in the area under the DVH curves (DVH AUC) for both the target and the five OARs. The decreasing solid lines represent the area change of the five OARs. We observe that the DVH AUC of the target continually increases, while the DVH AUC of the OARs decreases accordingly, which demonstrates the efficiency of the proposed model.

Figure 6 depicts the treatment planning process. The heat maps in the top panel represent the changes in dose distribution, while the corresponding DVH curves are depicted in the bottom panel at three stages: the initial step (left), step 50 (middle), and step 100 (right). The quality of the plans can be assessed based on the dose distributions of each structure, including the PTV and the five OARs. The bottom panel demonstrates the corresponding DVHs of each structure. For instance, the bottom left panel visualizes the initial DVHs in solid lines, while the top left represents the corresponding initial dose distribution, with segmentation curves delineated by the clinician. Changes in dose distribution in these areas help understand the dynamic planning process. The bottom middle and bottom right panels show the results of steps 50 and 100, respectively. At step 50, the DVH changes are compared by plotting both the initial step results (solid lines) and step 50 results (dashed lines). It is observed that the PTV and OAR curves shift at step 50, indicating a change in the dose distribution. At step 100, the dose level gap between PTV and OARs becomes more pronounced. The overall dose distribution in the top right panel maintains the expected distribution as labeled by the clinician. The result indicates that 95% volume of PTV would receive the prescription dose at stage three.

**Figure 6:**
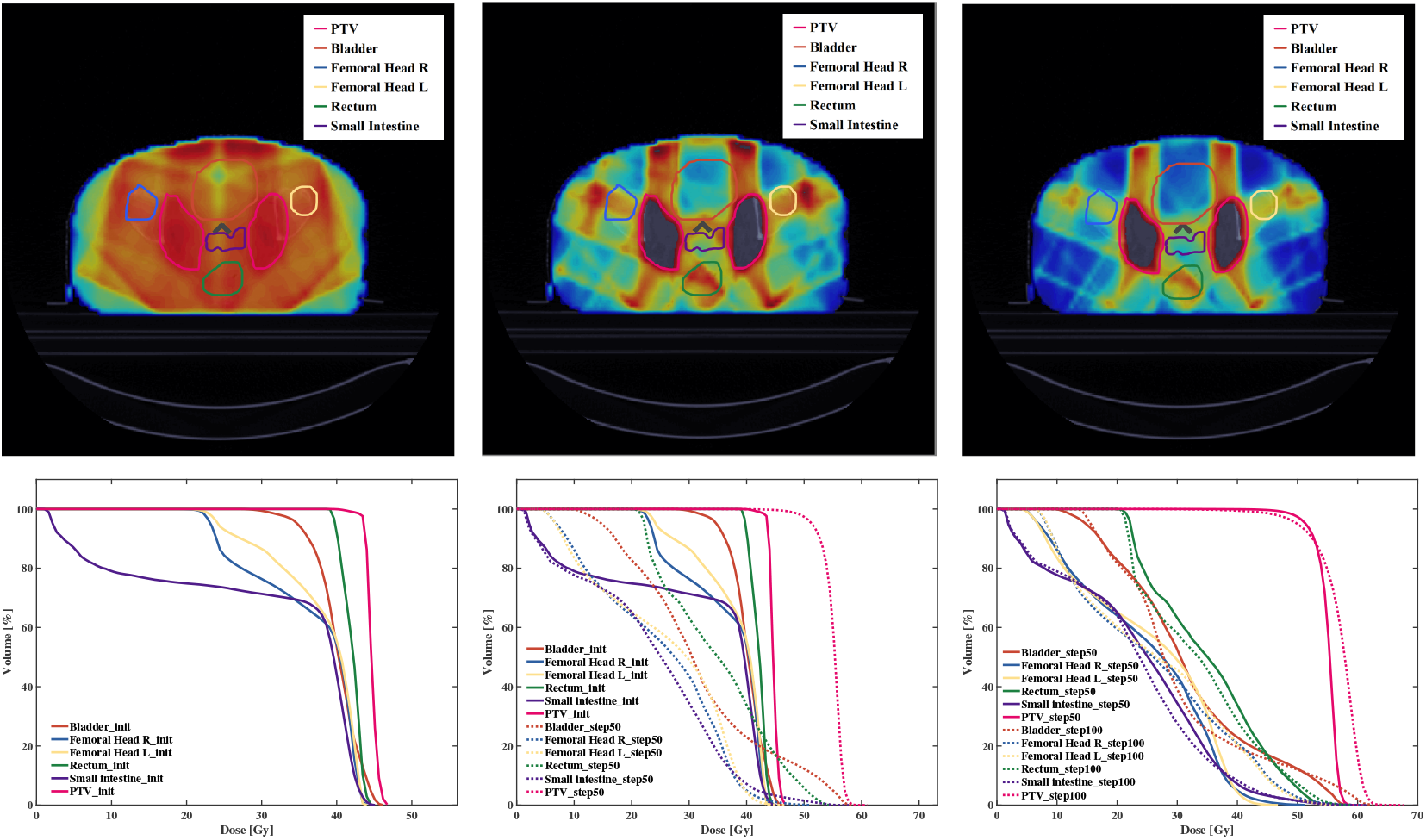
Variations of DVHs and dose distributions.

### 3.2 Clinical Results

Our experiment covers 12 structures, including both targets and OARs. We have omitted the results of the 6 PTV rings and reported only the results of the other 6 most important structures. Table 1 illustrates the key DVH parameters (with standard deviation) that clinicians most interested in during planning process of the 6 structures including one PTV and five OARs. For PTV, we evaluate D_98_/ D_95_ (the minimum absorbed dose that covers 98%/95% of the volume of the region of interest) and mean dose (D_mean_). The D_98_ of the Planning Target Volume (PTV) increased from the initial 44.91 Gy to 51.12 Gy, and the D_95_ increased from the initial 45.24 Gy to 51.68 Gy. It is noteworthy that the final D_95_ already meets the clinician standard 50.40 Gy.

**Table 1:** Summary of Interested DVH Parameters Changes (with standard deviation) to the PTV, and OARs.

| ROI | Parameters | Init | Step50 | Step100 |
| --- | --- | --- | --- | --- |
| PTV | D<sub>98</sub>(Gy) | 44.91±0.65 | 51.12±0.31 | 50.42±0.43 |
|  | D<sub>95</sub>(Gy) | 45.24±0.98 | 51.68±0.18 | 51.35±0.21 |
|  | D<sub>2</sub>(Gy) | 53.48±0.32 | 59.12±0.30 | 61.32±0.43 |
|  | D<sub>mean</sub>(Gy) | 46.08±0.37 | 56.34±0.62 | 57.71±0.15 |
| Bladder | V<sub>30</sub>(%) | 99.73±0.11 | 56.14±2.32 | 44.22±2.32 |
|  | V<sub>40</sub>(%) | 60.44±3.91 | 21.43±7.51 | 17.59±9.93 |
|  | V<sub>50</sub>(%) | 0 | 11.23±1.39 | 11.32±2.57 |
|  | D<sub>mean</sub>(Gy) | 42.89±1.23 | 33.24±1.42 | 30.80±1.72 |
| Rectum | V<sub>30</sub>(%) | 99.90±0.10 | 59.49±3.26 | 56.21±0.35 |
|  | V<sub>40</sub>(%) | 78.23±8.14 | 28.34±3.71 | 29.74±0.82 |
|  | V<sub>50</sub>(%) | 0 | 9.01±8.42 | 10.12±7.60 |
|  | D<sub>mean</sub>(Gy) | 43.14±1.26 | 31.53±1.37 | 33.48±0.84 |
| Small intestine | V<sub>30</sub>(%) | 45.91±7.38 | 20.70±2.14 | 17.84±6.68 |
|  | V<sub>40</sub>(%) | 17.34±6.01 | 5.54±2.90 | 0.54±0.11 |
|  | V<sub>50</sub>(%) | 0 | 2.66±0.70 | 2.32±0.41 |
|  | D<sub>mean</sub>(Gy) | 21.19±0.87 | 15.90±1.27 | 15.32±1.03 |
| Femoral Head R | V<sub>30</sub>(%) | 97.10±0.41 | 59.59±2.32 | 60.62±0.31 |
|  | V<sub>40</sub>(%) | 40.69±7.54 | 5.73±2.43 | 19.96±0.23 |
|  | V<sub>50</sub>(%) | 0 | 0 | 0.43±0.09 |
|  | D<sub>mean</sub>(Gy) | 39.51±0.71 | 29.65±0.73 | 30.57±0.73 |
| Femoral Head L | V<sub>30</sub>(%) | 90.57±5.42 | 62.02±0.88 | 63.31±7.60 |
|  | V<sub>40</sub>(%) | 46.34±6.35 | 9.42±8.50 | 25.65±6.31 |
|  | V<sub>50</sub>(%) | 0 | 0 | 0.21±0.13 |
|  | D<sub>mean</sub>(Gy) | 39.82±1.39 | 31.16±1.33 | 32.67±1.43 |

For the Organs at Risk (OARs), most DVH parameters, other than V_50_ at step 50, are significantly lower than the initial values. After 100 steps of adjustment, some DVH parameters still showed a certain improvement. Although the V_50_ of most organs has slightly increased, it remains within clinical requirements. For example, compared with the results at step 50, the initial V_30_ and V_40_ of the bladder, rectum, small intestine, and the right and left femoral heads decreased by 43.71% and 64.54%, 40.45% and 63.77%, 54.91% and 68.05%, 38.63% and 85.92%, 31.52% and 79.67%, respectively. Compared with the results of step 100, V_30_ and V_40_ in step 50 of the bladder and small intestine decreased by 21.23% and 17.91%, 13.81% and 90.25%, respectively.The V_30_ of rectum decreased by 5.51% during the adjustment from step 50 to step 100.

In comparison with the results at initial step, the V_30_ and V_40_, and mean dose of all OARs at step 50 were reduced. More specifically, in comparison with the initial mean dose, the single mean dose of the bladder, rectum, small intestine, and the right and left femoral heads at step 50 were reduced by 9.65 Gy (−22.50%), 11.61 Gy (−26.91%), 5.29 Gy (−24.96%), 9.86 Gy (−24.96%), and 8.66 Gy (−21.75%), respectively.

## 4 Discussion

The proposed automatic treatment planning framework was developed to mimic the behavior of human planners. It addresses two key challenges in automatic treatment planning. The first one is we broke through the restriction that the number of organs to be planned must be the same for different patients. In radiotherapy planning, even for patients with the same type of cancer, the structures (organs) of radiotherapy to be considered will be different. However, traditional DRL-based models design an exclusive sub-network for each organ to be planned, which implies the model to be consistent with the training patient in the test patient, severely reducing the scalability of the model [15, 10, 7]. To improve the scalability, we only use one network that is shared by all structures in FatPIN. During training, the functional features of DVH curves of different structures are fed into the network sequentially, and the output rewards of each organ are accumulated in one round of iterations before the gradient is passed back. In this way, the proposed model can be applied to real clinical scenarios where the number of organs planned for radiotherapy need not be the same for different patients. That is, personal planning protocols can be implemented for different patients.

Secondly, to enhance the efficiency of model learning, we introduced modifications to the input and output of FatPIN. On the input side, we devised a functional data embedding layer to extract features from the DVH curves. The existing models choose to process DVH curves directly with convolution [7, 10, 16], which ignore the functional data characteristics of DVH itself, making the signal-to-noise ratio of the processed features low and leading to low model learning efficiency. Hence,the proposed functional data embedding layer enables FatPIN to provide efficient computation for complex tumor patients. Regarding the treatment planning parameters to be output, we posited that they follows a normal distribution, rather then the traditional treatment of considering it as discrete classification problems. Consequently, we link the final output features with the mean and variance of a normal distribution. Continuous parameter adjustment was then obtained through sampling techniques. It is noteworthy that we can obtain the adjustment values with high confidence by constraining the variance.

Currently, automated radiotherapy planning algorithms proposed in the existing literature are far from clinical needs and are generally considered to be solved under a simplified framework only. For example, some of the existing methods require additional human intervention to narrow the state-action space [10]. And in [10], the authors only evaluate the case of one PTV and two OARs. This is because as the number of organs to be considered increases, the process of solving the optimization problem becomes more complex or even infeasible. In contrast, our algorithm makes the computational process efficient due to the two aforementioned strategies, thus simplifying the solution process. For example, the results demonstrate in Figure 5, Figure 6 and Table 1 are the treatment planning for cervical carcinoma patients which involve 12 areas. In the example of Figure 6, the results of 12 organs are actually processed, and only the results of the most critical 6 of them are kept in order to facilitate the presentation of the DVH curves. In Table 1, we give the results of a patient with 12 structures to be considered during treatment planning.

There are several limitations of our work. First, we know that radiotherapy planners are able to make planning across tumor categories. Although we have designed the organ-sharing TPPs network for patients from a same kind of cancer but with different number of structures, it is poorly trained in cases with different tumors. Second, the computational speed of our system is not fully satisfactory, as it takes 3 to 4 hours for a single test, which is comparable to the time taken by a human planner. This undermines the advantage of employing machine automation. The most time-consuming computational module is the interaction with the environment matRad [14], a MatLab-based TPS. During this interaction, a new optimization problem based on the action selection results predicted by DRL, needs to be addressed. Our model then transmits the constraint parameters of this new optimization problem to matRad to perform the next step of the optimization solution.

## 5 Conclusion

We present FatPIN, an organ-sharing network specifically devised for automatic radiotherapy treatment planning. FatPIN integrates a functional embedding layer to extract features from DVH curves and introduces a novel methodology for learning dose adjustment distribution, eschewing the reliance on predefined discrete adjustment steps. Experimental assessments carried out on cervical cancer cases showcase notable enhancements in patient metrics facilitated by the proposed network, thereby validating its direct utility in clinical contexts.

## Data Availability

All data produced in the present study are available upon reasonable request to the authors

